# Adverse childhood experiences and recent negative events activate immune and growth factor pathways, which are associated with first episode major depression and suicidal behaviours

**DOI:** 10.1101/2023.06.19.23291597

**Authors:** Abbas F. Almulla, Ali Abbas Abo Algon, Michael Maes

## Abstract

**Background:** Adverse Childhood Experiences (ACEs) and Negative Life Events (NLEs) may activate immune-inflammatory pathways, which play a role in the onset of Major Depressive Disorder and its severe phenotype Major Dysmood disorder (MDMD).

**Objectives:** To assess if elevated ACEs and NLEs in first episode (FE)-MDMD predict activation of the immune-inflammatory response system (IRS), chemokines, and growth factors that participate in the pathophysiology of MDMD.

**Methods:** This research assessed the effects of ACEs and NLEs on forty-eight cytokines/chemokines/growth factors, in 71 FE-MDMD patients and forty heathy controls.

**Results:** ACEs are highly significantly associated with the classical M1 macrophage, T helper (Th)-1, Th-1 polarization, IRS, and neurotoxicity immune profiles, and not with the alternative M2, and Th-2 immune profiles. There are highly significant correlations between ACEs and NLEs and different cytokines/chemokines/growth factors, especially with interleukin (IL)-16, CCL27, stem cell growth factor, and platelet-derived growth factor. Partial Least Squares analysis showed that 62.3% of the variance in the depression phenome (based on severity of depression, anxiety and suicidal behaviors) was explained by the regression on IL-4 (p=0.001, inversely), the sum of ACEs + NLEs (p<0.0001), and a vector extracted from 10 cytokines/chemokines/growth factors (p<0.0001; both positively associated). The latter partially mediated (p<0.0001) the effects of ACE + NLEs on the depression phenome.

**Conclusions:** Part of the effects of ACEs and NLEs on the depression phenome is mediated via activation of immune and growth factor networks. These pathways have a stronger impact in subjects with lowered activities of the compensatory immune-regulatory system.

## Introduction

Several studies suggest that major depressive disorder (MDD) may be classified as an immune-oxidative stress disease. This is supported by evidence indicating that MDD is accompanied by an activated immune response system (IRS) with upregulated T helper (Th)-1 cells, and increased levels of interleukin-2 (IL-2), soluble IL-2 receptor (sIL-2R), and interferon (IFN)-γ (Maes and Carvalho 2018). Additionally, M1 macrophage activities are indicated by high levels of IL-1β, IL-6, and tumor necrosis factor (TNF)-α (Maes, Bosmans et al. 1991, Maes, Scharpé et al. 1994, Maes, Smith et al. 1995, Mikova, Yakimova et al. 2001). In addition, it has been observed that individuals with MDD exhibit an augmented compensatory immune-regulatory system (CIRS) that functions as a counter-immune regulator (Maes, Berk et al. 2012). This is evidenced by elevated levels of IL-4 and IL-10, which are indicative of activated Th-2 and T regulatory (T reg) cell activation, respectively (Haapakoski, Mathieu et al. 2015, Köhler, Freitas et al. 2017, Maes and Carvalho 2018).

Maes and colleagues have utilized a novel methodology that leverages machine learning techniques to demonstrate the existence of two distinct phenotypes of MDD, specifically Major Dysmood Disorder (MDMD) and Simple DMD (SDMD) (Maes, Moraes et al. 2021, Maes, Rachayon et al. 2022). MDMD is characterized by elevated levels of depression, anxiety, fatigue, suicidal behaviors (SB), and physio-somatic symptoms. The research indicates heightened cognitive impairments and disabilities, as well as anomalies in autoimmunity, oxidative stress, antioxidant defenses, and gut dysbiosis in MDMD, which are not observed in SDMD (Maes, Moraes et al. 2021, Simeonova, Stoyanov et al. 2021, Maes 2022, Maes, Rachayon et al. 2022, Maes, Vasupanrajit et al. 2023). Upon stimulation, MDMD exhibited a significantly elevated production of M1, Th-1, Th-2, Th-17, Treg, IRS, CIRS, and growth factor production in comparison to the control group. The aforementioned anomalies were absent in SDMD (Maes, Rachayon et al. 2022). Almulla et al. (2023) have reported that the first episode (FE)-MDMD is concomitant with the activation of IRS and CIRS, as well as an elevation in immune-related neurotoxicity (Almulla, Ali Abbas Abo et al. 2023). The study conducted by Almulla et al. (2023) revealed that IL-16 was identified as the primary cytokine biomarker linked to FE-MDMD (Almulla, Ali Abbas Abo et al. 2023).

The likelihood of developing MDD, anxiety disorders, and suicidal behaviors in adulthood is greater for individuals who experienced multiple traumatic events during their childhood (Agnew-Blais and Danese 2016, Maes, Moraes et al. 2019). An increasing body of literature indicates that Adverse Childhood Experiences (ACEs), encompassing physical, emotional, and sexual abuse, as well as Negative Life Events (NLEs) such as bereavement and severe illness or injury, are significantly implicated in the etiology of MDD (Paykel, Emms et al. 1980, Kendler, Karkowski et al. 1999, Kraaij, Arensman et al. 2002, Maes, Congio et al. 2018, Abe, Sirichokchatchawan et al. 2022, Maes, Rachayon et al. 2022). In the present context, it has been observed through the utilization of Research and Diagnostic Algorithmic Rules (RADAR) that the occurrence of first episode MDD and current suicidal behaviors are predominantly linked to the cumulative impact of ACEs and NLEs (Abe, Sirichokchatchawan et al. 2022, Maes and Almulla 2023).

The presence of ACEs and severe socioeconomic deprivation have been found to be significantly correlated with heightened inflammatory responses during adulthood, as well as various neurological and autoimmune disorders (Maes, Moraes et al. 2019). Maes et al. (2022) observed a correlation between ACEs and the manifestation of affective disorders, immune-related neurotoxicity, and growth factor profiles. This was determined through the analysis of various cytokines, chemokines, and growth factors present in the culture supernatant of stimulated whole blood. However, there is a lack of published research investigating the influence of ACEs and NLEs on serum cytokines, chemokines, and growth factors in patients with FE-MDMD.

Hence, the current study aimed to delineate the potential correlation between ACEs and NLEs and immune profiles among individuals diagnosed with FE-MDMD. Using a precision nomothetic methodology (Maes 2022, Maes and Stoyanov 2022), the specific hypothesis is that ACEs and NLEs can predict the abnormal immune-inflammatory pathways that are implicated in the pathophysiology of FE-MDMD.

## Materials and methods

### Participants

A senior psychiatrist in Alhakiem Hospital, Najaf, Iraq, identified and enlisted 71 individuals with FE-MDD in accordance with the Diagnostic and Statistical Manual of Mental Disorders (DSM-5) (American Psychiatric Association 2013) criteria. Additionally, all FE-MDD patients fulfilled the MDMD diagnostic requirements (Maes, Vasupanrajit et al. 2023). The MDMD diagnosis can be made using the Hamilton Depression Rating Scale (HAMD) (Hamilton 1960), Hamilton Anxiety Rating Scale (HAMA) (Hamilton 1959), suicidal behaviors, and recurrence of disease scores (Maes and Almulla 2023). When both the HAMD and HAMA scores are greater than twenty-two, it can also be employed. All the FE-MDMD patients were in the acute stage of the illness and lacked any indicators of full or partial remission. The same senior psychiatrist also recruited forty healthy individuals from the same catchment area, including hospital personnel and acquaintances of the patients, to serve as the control group. The entire group was enlisted between October 2021 and March 2022.

Participants who met the DSM-5 criteria for any of the following conditions were omitted: dysthymia (except double depression), schizophrenia, schizoaffective disorder, bipolar disorder, autism spectrum disorders, substance use disorders (nicotine dependence was the exception), post-traumatic stress disorder, psycho-organic disorders, generalized anxiety disorder, and obsessive-compulsive disorder. We also took great care when selecting the control group. Controls with a history of major depression or dysthymia or a family history of depression, mania, psychosis, substance use disorder, or suicide were not considered. Individuals with chronic liver or kidney disease, pregnant and breastfeeding women, and those suffering from stroke, multiple sclerosis, Parkinson’s disease, or Alzheimer’s disease were excluded. The same went for (auto)immune diseases like psoriasis, rheumatoid arthritis, inflammatory bowel disease, cancer, type 1 diabetes, and scleroderma. We also avoided the participation of individuals who had suffered from an acute COVID-19 infection, severe or critical COVID-19 disease, Long COVID, or had a COVID-19 infection within 6 months prior to enrollment. Finally, we did not include individuals being treated with immunosuppressive or immunomodulatory medications or taking therapeutic doses of antioxidants or omega-3 supplements.

Before taking part in the study, each participant, or, where appropriate, their parents or legal guardians, gave their written informed consent. The College of Medical Technology Ethics Committee of the Islamic University of Najaf in Iraq approved this study (Document No. 18/2021). The research was conducted strictly following local Iraqi and international ethical and privacy regulations. This includes but is not limited to the principles outlined in the World Medical Association’s Declaration of Helsinki, the Belmont Report, the CIOMS Guideline, and the International Conference on Harmonization of Good Clinical Practice. Our institutional review board is committed to maintaining the highest standards, strictly adhering to the International Guideline for Human Research Safety (ICH-GCP).

### Clinical assessments

A senior psychiatrist employed a semi-structured interview to gather sociodemographic, clinical, and psychological information. To define the severity of depression and anxiety, respectively, the total scores on the HAMD (Hamilton 1960) and HAMA (Hamilton 1959) were used. We employed the total and “pure” (i.e., devoid of physio-somatic symptoms) HAMD depression and HAMA anxiety scores in the current investigation. The former was conceived of as the sum of a depressed state, guilt feelings, suicide thoughts, and interest loss. The pure anxiety score was computed as the sum of anxious mood, tension, fears, and anxious behavior during the interview. Two items from the Columbia Suicide Severity Rating Scale (C-SSRS) were used to evaluate suicidal behaviors: a) the number of suicide attempts within the previous year, and b) the frequency of suicidal thinking within the previous three months (Posner, Brown et al. 2011). We calculated a composite score of last year’s suicidal behaviors (SB), weighted in z units, by adding the z scores for the HAMD suicide item, the number of suicide attempts, and the frequency of suicidal ideation.

The ACE Questionnaire was employed to determine the severity of ACEs (Felitti, Anda et al. 1998). This scale comprises 28 items, including scoring of 10 domains, namely (1) mental trauma, (2) physical trauma, (3) sexual abuse, (4) mental neglect, (5) physical neglect, (6) witnessing a mother being abused (domestic violence), (7) family member with drug abuse, (8) family member with depression/mental illness, (9) losing a parent to separation, death, divorce, and (10) a family member who is in prison. We computed different ACE scores: total sum of all ACEs (total ACE), sum of physical trauma + mental neglect + family member with drug abuse (ACE247) and sum of ACE247 + a family member who is in prison (ACE24710). In addition, we used the negative life events (NLEs) scale to evaluate NLEs that occurred the year prior to inclusion in our study (Buri 2018). This scale involved 16 items, and in the present study we used the following items: serious accident, death of a family member or close friend, divorce or separation, not being able to get a job, lost job, alcohol-related problems, drug-related problems, seeing fights or people beaten up, abuse or violent crime, trouble with the police, gambling problem, member of the family sent to jail, overcrowding at home, discrimination/racism, vandalism or malicious property damage. Consequently, we computed the sum of total ACE + the presence of one or more NLEs, labeled ACE+NLEs. We split the study sample into three groups based on the number of ACE+NLEs and formed three subgroups: those without any ACE+NLEs (labeled ACE+NLEs 0; n=25), those with 1 or 2 ACE+NLEs (labeled ACE+NLEe 1-2; n=53), and those with three or more ACE+NLEs (labeled ACE+NLEs ≥3; n=33). Participants’ body mass index (BMI) was calculated by dividing their kilogram body weight by their square meter height. Tobacco use disorder was assessed using DSM-5 criteria.

### Biochemical assays

We used a disposable syringe and serum tubes to collect 5 mL of fasting venous blood from each participant in the trial between 8:00 and 11:00 in the morning. The serum and blood cells were separated after the blood was centrifuged at 35,000 revolutions per minute (rpm). The serum was then put into small Eppendorf tubes kept at –80 C, until required for the biomarker assays. The levels of forty-eight cytokines/chemokines/growth factors were measured in the serum of all participants using Bio-Plex Multiplex Immunoassay kits (Bio-Rad Laboratories Inc., Hercules, USA). The kits use a fluorescence-based method to measure the levels of these proteins in blood serum. The fluorescence intensity (FI) of each protein was measured, and the blank subtracted values were used in the study (Rachayon, Jirakran et al. 2022). We followed a procedure including several steps detailed in our previous study (Almulla, Ali Abbas Abo et al. 2023). **Electronic supplementary file (ESF)**, Table 1, itemizes all the analytes identified in our research and it also provides their corresponding gene IDs and alternate names. The intra-assay CV values for all analytes were below 11.0%. To determine the concentrations of the analytes, we referred to the manufacturer-supplied standards. We then calculated the out-of-range (OOR) concentration rate, i.e., the proportion of concentrations below the minimum detectable level. In our statistical analysis, we discarded any cytokines/chemokines/growth factors whose concentrations were, in over 80% of the cases, lower than the lowest out-of-range (OOR) value. Consequently, our study did not include IFN-α2, IL-3, IL-7, and IL-12p40. When constructing our composite scores, we considered analytes that had measurable levels between 20% and 40% as prevalence rates (or dummy variables). As such, we calculated different immune profiles (as described in the introduction and ESF, Table 2), which included all analytes, unless their detectable concentrations were less than 20% (Rachayon, Jirakran et al. 2022, Thisayakorn, Thipakorn et al. 2022, Almulla, Ali Abbas Abo et al. 2023). The primary outcome data in this study are the immunological profiles constructed using z-unit-weighted composite scores of several cytokines, chemokines, and growth factors. The variables utilized to create M1, M2, M1/M2 (z M1 – z M2), Th1, Th2, Th1/Th2 (z Th-1 – z Th-2), IRS, CIRS, IRS/CIRS (z IRS – z CIRS), and neurotoxicity profiles are listed in ESF, Table 2. We also analyzed the individual cytokines/chemokines/growth factors if the immune profiles revealed significant differences between the study groups.

**Table 1:** Sociodemographic and clinical characteristics in three study groups based on number of adverse childhood experiences (ACEs) and negative life events (NLEs).

| Variables | ACE+NLEs 0<br>(n=25)<sup>A</sup> | ACE+NLEs 1-2<br>(n=53)<sup>B</sup> | ACE+NLEs ≥3<br>(n=33)<sup>C</sup> | F | df | p |
| --- | --- | --- | --- | --- | --- | --- |
| Age (Years) | 31.48(7.44) | 31.83(8.81) | 30.94(8.76) | 0.111 | 2/108 | 0.895 |
| BMI (Kg/m<sup>2</sup>) | 26.33(4.03) | 24.70(3.61) | 24.10(3.78) | 2.614 | 2/108 | 0.078 |
| Educated (Y/N) | 19/6 | 41/12 | 23/10 | 1.638 | 2 | 0.441 |
| Sex (M/F) | 12/13 | 22/31 | 13/20 | 0.460 | 2 | 0.794 |
| Marital State (Y/N) | 14/11 | 22/31 | 11/22 | 3.022 | 2 | 0.221 |
| Smoking (Y/N) | 10/15 | 18/35 | 8/25 | 1.720 | 2 | 0.423 |
| Previous COVID infection (Y/N) | 11/14 | 18/35 | 15/18 | 1.379 | 2 | 0.502 |
| ACE247 | 0<sup>A,C</sup> | 0.641(0.623)<sup>B,C</sup> | 1.697(0.728)<sup>A,B</sup> | KWT |  | <0.001 |
| ACE24710 | 0<sup>A,C</sup> | 0.660(0.618)<sup>B,C</sup> | 1.878(0.819)<sup>A,B</sup> | KWT |  | <0.001 |
| ACE total | 0<sup>A,C</sup> | 1.06(0.456)<sup>B,C</sup> | 2.64(0.783)<sup>B,C</sup> | KWT |  | <0.001 |
| ACE + NLEs | 0<sup>A,C</sup> | 1.41(0.497)<sup>B,C</sup> | 3.42(0.662)<sup>B,C</sup> | KWT |  | <0.001 |
| Pure Depression | 1.40(3.41)<sup>B,C</sup> | 7.24(4.79)<sup>A,C</sup> | 11.03(1.44)<sup>A,B</sup> | 46.30 | 2/108 | <0.001 |
| Pure Anxiety | 1.84(2.74)<sup>B,C</sup> | 6.43(4.06)<sup>A,C</sup> | 9.15(1.73)<sup>A,B</sup> | 36.37 | 2/108 | <0.001 |
| Suicidal Behaviors (z score) | -1.018(0.563)<sup>B,C</sup> | 0.085(1.021)<sup>A,C</sup> | 0.633(0.500)<sup>A,B</sup> | 30.29 | 2/108 | <0.001 |
| Phenome (z score) | -1.12(0.612)<sup>B,C</sup> | 0.069(0.969)<sup>A,C</sup> | 0.741(0.250)<sup>A,B</sup> | 45.21 | 2/108 | <0.001 |

The study groups were formed using the sum of total ACE and NLEs (ACE+NLEs), namely those without any ACE+NLEs (ACE+NLEs 0), those with 1 or 2 ACE+NLEs (ACE+NLEs 1-2) and those with three or more ACE+NLEs (ACE+NLEs $\geq 3$ ).
ACE247: sum of the ACE Questionnaire items Q2 + Q4 + Q7; ACE24710: sum of items Q2 + Q4 + Q7 + Q10; total ACE: sum of all ACEs.
Results are shown as mean ( ± SD). F: results of analysis of variance; $X^2$ : analysis of contingency tables, KWT: Kruskal-Wallis test; BMI: Body mass Index, Kg: Kilograms, m: meter, Y: Yes, N: No, M: Male, F: Female.
Pure depression and anxiety: depressive and anxiety items of the Hamilton Depression and Anxiety Rating Scale (thus without any of the physiosomatic symptoms); phenome: latent vector extracted from the Hamilton Depression and Anxiety Rating Scale and suicidal behaviors scores.

**Table 2:** Differences in three study groups based on number of adverse childhood experiences (ACEs) and negative life events (NLEs).

| Variables (z score) | ACE+NLEs 0<br>(n=25)<sup>A</sup> | ACE+NLEs 1-2<br>(n=53)<sup>B</sup> | ACE+NLEs $\geq 3$<br>(n=33)<sup>C</sup> | F | df | p-value |
| --- | --- | --- | --- | --- | --- | --- |
| M1-classical | -0.496(0.202)<sup>B,C</sup> | 0.048(0.138)<sup>A</sup> | 0.275(0.169)<sup>A</sup> | 4.39 | 2/92 | 0.015 |
| M2-alternative | -0.261(0.211) | 0.011(0.144) | 0.167(0.176) | 1.23 | 2/92 | 0.298 |
| zM1 - z M2 | -0.235(0.133) | 0.038(0.091) | 0.108(0.111) | 2.11 | 2/92 | 0.127 |
| Th-1 | -0.411(0.201)<sup>C</sup> | -0.006(0.137) | 0.296(0.168)<sup>A</sup> | 3.66 | 2/92 | 0.030 |
| Th-2 | -0.199(0.209) | 0.019(0.143) | 0.111(0.175) | 0.67 | 2/92 | 0.516 |
| zTh1 - zTh2 | -0.212(0.114)<sup>C</sup> | -0.024(0.078) | 0.185(0.095)<sup>A</sup> | 3.68 | 2/92 | 0.029 |
| Th-17 | -0.390(0.209) | 0.066(0.143) | 0.174(0.175) | 2.34 | 2/92 | 0.102 |
| IRS | -0.456(0.203)<sup>C</sup> | 0.017(0.138) | 0.294(0.169)<sup>A</sup> | 4.05 | 2/92 | 0.021 |
| CIRS | -0.317(0.213) | 0.031(0.145) | 0.176(0.178) | 1.62 | 2/92 | 0.203 |
| zIRS - zCIRS | -0.330(0.211) | -0.033(0.144) | 0.280(0.176) | 2.52 | 2/92 | 0.086 |
| Neurotoxicity | -0.441(0.205)<sup>C</sup> | 0.014(0.140) | 0.287(0.171)<sup>A</sup> | 3.72 | 2/92 | 0.028 |

The study groups were formed using the sum of total ACE and NLEs (ACE+NLEs), namely those without any ACE+NLEs (ACE+NLEs 0), those with 1 or 2 ACE+NLEs (ACE+NLEs 1-2) and those with three or more ACE+NLEs (ACE+NLEs $\geq 3$ ).
ACE247: sum of the ACE items Q2+Q4+Q7; ACE24710: sum of items Q2+Q4+Q7+Q10; total ACE: sum of all ACEs.
M: Macrophage, Th: T helper, IRS: Immune response system, CIRS: Compensatory Immune response system.

### Statistics

In the present research, all statistical analyses were executed utilizing IBM SPSS 28, Windows version. Continuous variables were compared across study groups by applying analysis of variance (ANOVA) or the Kruskal-Wallis test. Conversely, nominal variables were compared via contingency table analysis, specifically the Chi-square test. Furthermore, Pearson’s and point-biserial correlation coefficients were employed to examine the associations between scale variables or scale and binary variables. Manual multiple regression analysis was utilized to elucidate the influence of specific ACEs, NLEs, and additional demographic attributes on the immune profiles. Similarly, these analyses were employed to scrutinize the impact of predictor variables, specifically the immune profiles, ACEs, and NLEs, on the outcome variables, which include the depression phenome. Furthermore, we employed an automatic forward stepwise regression technique. This method established a threshold of p=0.05 for the inclusion of variables, and p=0.06 for their exclusion. This strategy assisted us in determining which variables ought to be incorporated into the final regression model and which should be excluded. We calculated the standardized coefficients, t-statistics, and exact p-values for each variable incorporated into the final regression models. We also ascertained the F statistics, their corresponding p-values, and the measure of effect size was determined using R^2^ or partial Eta squared. We scrutinized the data for potential multicollinearity and collinearity by employing specific measures. A tolerance limit of 0.25 was established, and a variance inflation factor threshold exceeding 4 was set. Furthermore, we utilized the condition index and variance proportions extracted from the collinearity diagnostics table for this analysis. The presence of heteroskedasticity was confirmed by implementing the White test and the modified version of the Breusch-Pagan test. All the analyses above were executed employing two-tailed tests. A p-value (alpha level) of 0.05 or lower was deemed to represent statistical significance. We opted not to implement False Discovery Rate (FDR) p-value corrections, given the interconnected nature of all cytokines, chemokines, and growth factors within the immune (cytokines and chemokines) and growth factor networks (Maes, Plaimas et al. 2021, Rachayon, Jirakran et al. 2022). When required, we employed transformations, including logarithmic, square-root, and rank-based inversed normal (RINT), to attain a normal distribution for our data indicators.

To investigate the causal connections between ACE+NLEs, immunological profiles, and the depressive phenome, partial least squares (PLS) analysis was performed. The output variable was a latent vector that was extracted from the pure HAMD and HAMA scores as well as the SB scores. ACE+NLEs were input as a single indicator. The immune profiles were added as possible mediators between ACE+NLEs and the phenome. Only when the outer and inner models satisfied the following predetermined quality criteria was a complete PLS analysis performed: a) the latent vectors display high construct and convergence validity, as indicated by Cronbach’s alpha > 0.7, composite reliability > 0.8, and average variance extracted (AVE) > 0.5; b) blindfolding shows that the construct’s cross-validated redundancy is sufficient; c) confirmatory tetrad analysis (CTA) shows that the latent vectors extracted from the indicators are not mis-specified as reflective models; d) the model fit is adequate with standardized root squared residual (SRMR) <0.08; and e) all loadings on the extracted latent vectors are >0.65 at p < 0.001. Consequently, we performed complete PLS-SEM pathway analysis (SmartPLS) with 5,000 bootstrap samples and estimated the path coefficients (with p-values), as well as specific indirect, total indirect (mediated) effects, and total effects, if the model quality data met the predetermined conditions. The findings of multiple regression studies that used PLS analysis to evaluate how ACE+NLEs affect immune activation indices were the main statistical analyses. A priori power analysis (G*Power 3.1.9.4) suggests a minimum sample size of 103 for a linear multiple regression analysis with an effect size of 0.111 (equivalent to approximately 10% explained variance), alpha=0.05, power=0.8, and 3 covariates.

## Results

### Socio-demographic data

**Table 1** shows the sociodemographic characteristics, clinical data, affective symptoms, and suicidal behaviors of the three study groups formed using the ACE and NLEs data. Our findings show that the total ACE, ACE247, ACE24710, and ACE+NLEs, the pure depression and anxiety symptoms, SB, and phenome were significantly different among three study groups and increased from ACE+NLE 0 to ACE+NLE 1-2 to ACE+NLE ≥3. We also found that age, BMI, education level, sex, marital status, smoking, and previous COVID-19 infection did not exhibit significant differences between the study groups.

### Differences in immune profiles between study groups

The findings presented in **Table 2** demonstrate that M1-classical was higher in the ACE+NLEs ≥3 and ACE+NLEs 1-2 group as compared with the ACE-NLEs 0 group. Th-1, zTh1 – zTh2, neurotoxicity, and IRS were significantly higher in the ACE-NLEs ≥3 group versus the ACE-NLEs 0 group. Conversely, no significant differences were observed in M2-alternative, Th-2, Th-17, zM1 – z M2, CIRS, and the zIRS – zCIRS ratio between the study groups.

The results presented in ESF, Tables 3 and 4 indicate significant differences in the levels of sIL-2R, IL-6, IL-8, IL-9, IL-16, IL-18, TNF-β, TRAIL, MIF, CCL2, CCL4, CCL5, CCL27, CXCL9, CXCL10, M-CSF, SCF-1, SCGFB, SDF, PDGFB, and HGF among the various study groups. Specifically, IL-16, CCL27, SCGFB, and PDGFB were significantly different among the three study groups and increased from ACE+NLEs 0 to ACE+NLEs 1-2 to ACE+NLEs ≥3. On the other hand, the remaining biomarkers showed significant differences in ACE+NLEs ≥3 versus controls.

**Table 3:** Correlation matrix between adverse childhood experiences (ACEs) and negative life events (NLEs), and affective symptoms, suicidal behaviors, and the phenome of depression.

| Variables | Total ACE | ACE24710 | NLE | zHAMD+zHAMA | Suicidal Behaviors | Phenome |
| --- | --- | --- | --- | --- | --- | --- |
| M1-classical | 0.313** | 0.268** | 0.098 | 0.417** | 0.404** | 0.427** |
| M2-alternative | 0.176 | 0.118 | 0.061 | 0.330** | 0.294** | 0.329** |
| zM1 - z M2 | 0.224* | 0.244* | 0.060 | 0.141 | 0.180 | 0.159 |
| Th-1 | 0.268** | 0.245* | 0.107 | 0.419** | 0.367** | 0.416** |
| Th-2 | 0.108 | 0.056 | 0.065 | 0.269** | 0.248** | 0.271** |
| zTh1 - zTh2 | 0.301** | 0.355** | 0.080 | 0.282** | 0.223** | 0.272** |
| Th-17 | 0.200 | 0.170 | 0.093 | 0.275** | 0.194** | 0.256* |
| IRS | 0.282** | 0.249* | 0.150 | 0.443** | 0.381** | 0.437** |
| CIRS | 0.201* | 0.148 | 0.132 | 0.396** | 0.289** | 0.373** |
| zIRS - zCIRS | 0.191 | 0.241* | 0.191 | 0.112** | 0.218* | 0.152 |
| Neurotoxicity | 0.272** | 0.230* | 0.150 | 0.402** | 0.332** | 0.392** |

\*p<0.05; \*\*p<0.01; M: Macrophage, Th: T helper, IRS: Immune response system, CIRS: Compensatory Immune response system. ACE24710: sum of the ACE Questionnaire items Q2 + Q4 + Q7 + Q10; total ACE: sum of all ACE items; zHAMD+zHAMA: z unit based composite score based on the Hamilton Depression and Anxiety Rating Scale scores. phenome: latent vector extracted from the Hamilton Depression (HAMD) and Anxiety (HAMA) Rating Scale and suicidal behaviors scores.

**Table 4:** Results of multiple regression analyses with affective symptoms and suicidal behaviors as dependent variables, and adverse childhood experiences (ACEs), negative life events (NLEs), and immune profiles as explanatory variables.

| Dependent Variables | Explanatory Variables | Coefficients of input variables |  |  | Model statistics |  |  |  |
| --- | --- | --- | --- | --- | --- | --- | --- | --- |
| | | $\beta$ | t | p | R<sup>2</sup> | F | df | p |
| #1. Pure Depression | **Model** |  |  |  | 0.538 | 35.748 | 3/92 | <0.001 |
|  | ACE+NLEs | 0.459 | 5.049 | <0.001 |  |  |  |  |
|  | IRS | 0.249 | 3.370 | 0.001 |  |  |  |  |
|  | ACEQ4 | 0.228 | 2.567 | 0.012 |  |  |  |  |
| #2. Pure Anxiety | **Model** |  |  |  | 0.494 | 45.357 | 2/93 | <0.001 |
|  | ACE+NLEs | 0.598 | 7.772 | <0.001 |  |  |  |  |
|  | IRS | 0.236 | 3.071 | 0.003 |  |  |  |  |
| #3. Suicidal Behavior | **Model** |  |  |  | 0.402 | 20.598 | 3/92 | <0.001 |
|  | ACE+NLEs | 0.319 | 3.085 | 0.003 |  |  |  |  |
|  | M1-classical | 0.257 | 3.042 | 0.003 |  |  |  |  |
|  | ACEQ4 | 0.250 | 2.467 | 0.015 |  |  |  |  |
| #4. Phenome | **Model** |  |  |  | 0.533 | 34.966 | 3/92 | <0.001 |
|  | ACE+NLEs | 0.432 | 4.723 | <0.001 |  |  |  |  |
|  | IRS | 0.267 | 3.594 | <0.001 |  |  |  |  |
|  | ACEQ4 | 0.241 | 2.701 | 0.008 |  |  |  |  |

Pure depression and pure anxiety: the sum of depressive and anxiety items of the Hamilton Depression and Anxiety Rating Scale, respectively (thus without any of the physio-somatic symptoms); phenome: latent vector extracted from the Hamilton Depression and Anxiety Rating Scale and suicidal behaviors scores; ACE+NLEs: sum of total ACE and NLEs; ACEQ4: item 4, emotional neglect; IRS: immune-inflammatory response system, M1: classical M1 macrophage.

### Intercorrelations

There were significant associations between total ACE (r=0.333, p<0.001), ACE247 (r=0.333, p<0.001), ACE24710 (r=0.303, p<0.001) and NLEs. Correlation analyses examined the associations between total ACE, ACE247, ACE20710, NLE, affective symptoms, SB, the phenome, and the various immune profiles. The outcomes of correlation analyses are presented in **Table 3**. Total ACE and ACE24710 were significantly associated with M1 classical, zM1 – zM2, Th-1, zTh1 – zTh2, IRS and neurotoxicity profiles. Furthermore. total ACE was correlated with CIRS, and ACE24710 with zIRS – zCIRS. There were no significant associations between NLEs and any of the immune profiles. The results indicate that M1-classical, M2-alternative, Th-1, Th-2, zTh1 – zTh2, neurotoxicity, IRS, and CIRS display significant positive correlations with affective symptoms, SB, and the depression phenome. Furthermore, the zIRS – zCIRS scores exhibit a significant positive correlation with SB, while Th-17 is significantly correlated with both affective symptoms and the phenome (positive associations). Moreover, the results of other correlation analyses (presented in ESF, Table 5) provide insights into the significant relationships between cytokines/chemokines/growth factors and the total ACEs, ACE24710, NLEs, and ACE+NLEs. **Figure 1** shows the partial regression of serum IL-16 on ACE+NLEs, and **Figure 2** shows the partial regression of serum PDGF on ACE+NLEs (both after adjusting for age, sex, and BMI).

**Figure 1:**
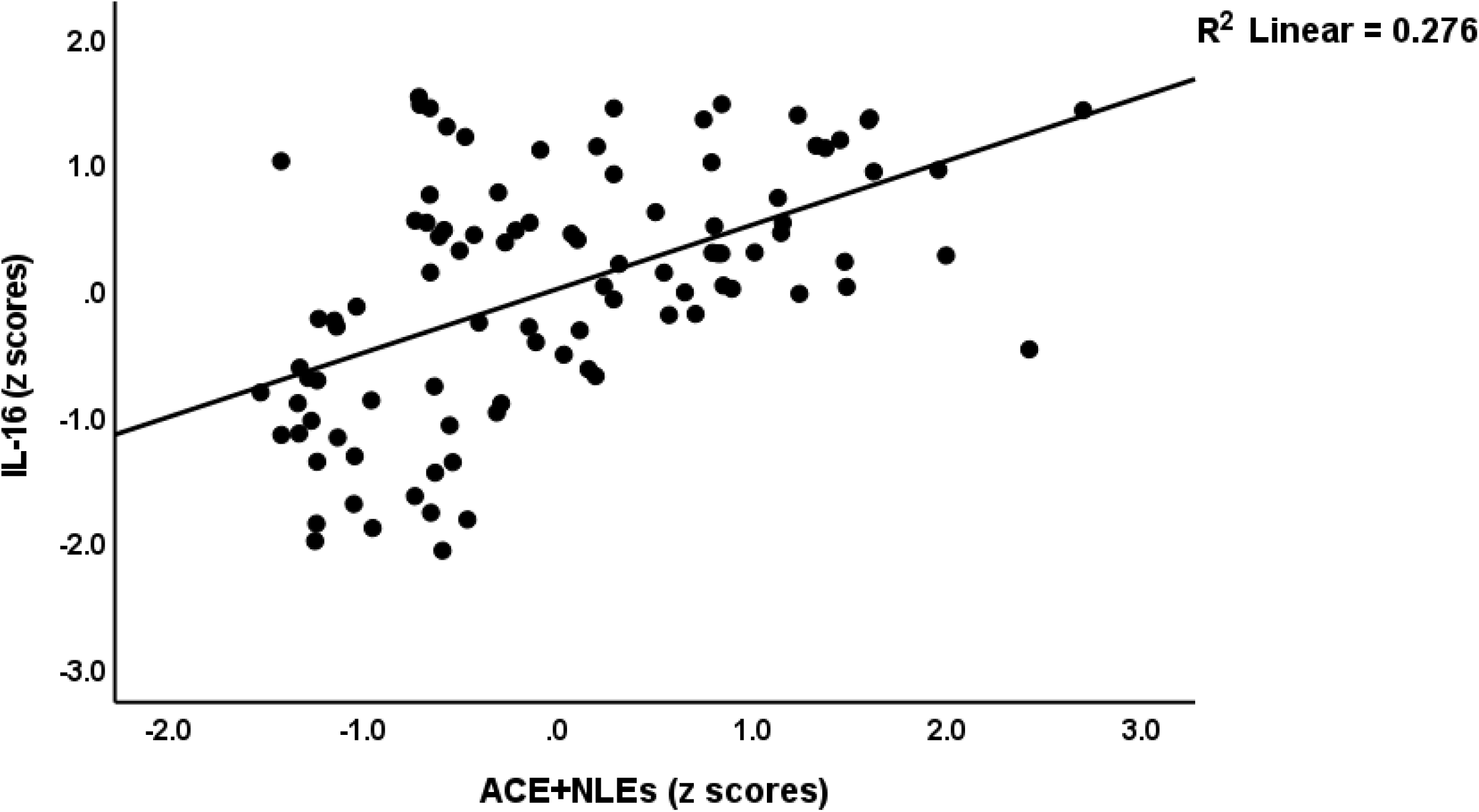
Partial regression of serum interleukin-16 (IL-16) on the sum of adverse childhood experiences (ACE) and negative life events (NLEs); t=5.89, p<0.001.

**Figure 2:**
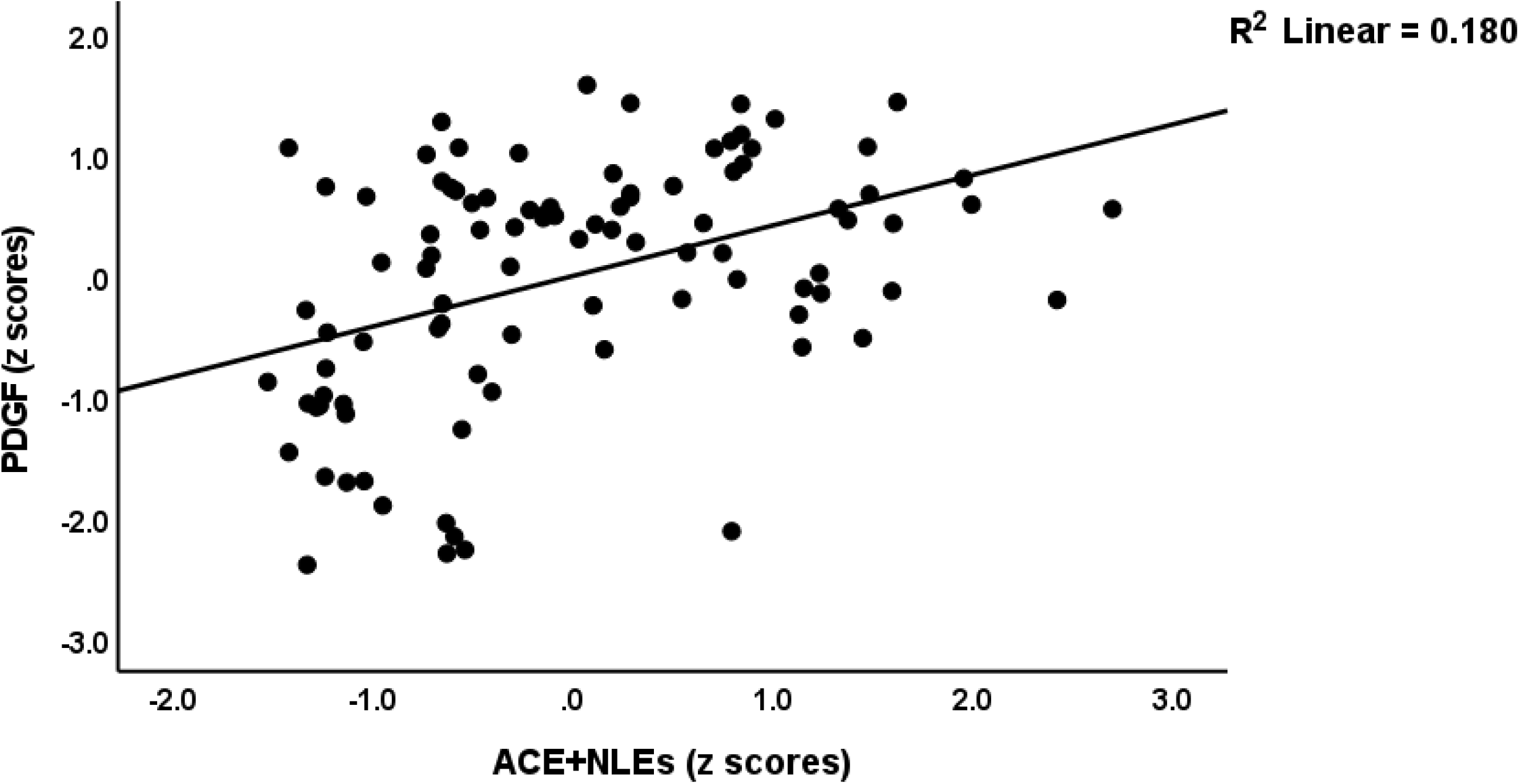
Partial regression of serum platelet derived growth factor (PDGF) on the sum of adverse childhood experiences (ACE) and negative life events (NLEs); t=4.47, p<0.001.

**Table 5:** Results of multiple regression analyses with affective symptoms and suicidal behaviors as dependent variables and adverse childhood experiences (ACEs) and negative life events (NLEs), cytokines, chemokines, and growth factors as explanatory variables.

| Dependent Variables | Explanatory Variables | Coefficients of input variables |  |  | Model statistics |  |  |  |
| --- | --- | --- | --- | --- | --- | --- | --- | --- |
| | | $\beta$ | t | p | R<sup>2</sup> | F | df | p |
| #1. Pure Depression | Model |  |  |  | 0.766 | 74.372 | 4/91 | <0.001 |
|  | IL-16 | 0.567 | 7.605 | <0.001 |  |  |  |  |
|  | CCL3 | -0.243 | -4.347 | <0.001 |  |  |  |  |
|  | ACE+NLEs | 0.270 | 4.334 | <0.001 |  |  |  |  |
|  | PDGF | 0.187 | 2.886 | 0.005 |  |  |  |  |
| #2. Pure Anxiety | Model |  |  |  | 0.730 | 61.415 | 4/91 | <0.001 |
|  | IL-16 | 0.492 | 5.739 | <0.001 |  |  |  |  |
|  | CCL3 | -0.240 | -4.067 | <0.001 |  |  |  |  |
|  | ACE+NLE | 0.277 | 4.118 | <0.001 |  |  |  |  |
|  | SCGF | 0.220 | 2.821 | 0.006 |  |  |  |  |
| #3. Suicidal behaviors | Model |  |  |  | 0.652 | 33.702 | 5/90 | <0.001 |
|  | IL-16 | 0.449 | 4.809 | <0.001 |  |  |  |  |
|  | SCGF | 0.298 | 3.350 | 0.001 |  |  |  |  |
|  | CCL3 | -0.282 | -4.180 | <0.001 |  |  |  |  |
|  | ACEQ4 | 0.171 | 2.448 | 0.016 |  |  |  |  |
|  | Gender | 0.142 | 2.235 | 0.028 |  |  |  |  |
| #4. Phenome | Model |  |  |  | 0.752 | 68.882 | 4/91 | <.001e |
|  | IL-16 | 0.567 | 7.224 | <0.001 |  |  |  |  |
|  | CCL3 | -0.240 | -4.294 | <0.001 |  |  |  |  |
|  | ACEQ4 | 0.224 | 3.835 | <0.001 |  |  |  |  |
|  | SCGF | 0.239 | 3.196 | 0.002 |  |  |  |  |

Pure depression and pure anxiety: the sum of depressive and anxiety items of the Hamilton Depression and Anxiety Rating Scale, respectively (thus without any of the physio-somatic symptoms); phenome: latent vector extracted from the Hamilton Depression and
Anxiety Rating Scale and suicidal behaviors scores; ACE+NLEs: sum of total ACE and NLEs; ACEQ4: item 4, emotional neglect; IL: interleukin; CCL3: C-C motif Chemokine ligand 3; SCGF: stem cell growth factor.

### Prediction of the depression phenome by ACEs, NLEs, and immune profiles

**Table 4**, regressions #1 and #4 show that large parts of the variances in pure depression (53.8%), and the phenome (53.3%) could be explained by ACE+NLEs, IRS and ACEQ4 (all positively associated). Regression #2 shows that ACE+NLEs and IRS (both positively associated) significantly predict 49.4% of the variance in pure anxiety. Regression #3 reveals that ACE+NLEs, M1-classical, and ACEQ4 (all positive associations) significantly predict 40.2% of the variance in SB.

**Table 5** shows the outcome of similar multiple regression analyses with clinical scores as dependent variables and ACEs, NLEs, and single cytokines/chemokines/growth factors as explanatory variables. Regression #1 indicates that an extensive part of the variance (76.6%) in the pure depression score could be significantly predicted by IL-16, ACE+NLEs, PDGFB (all positively associated) and CCL3 (inversely associated). Regression #2 displays that IL-16, ACE+NLEs, and SCGF (all positive associations) and CCL3 (negative association) predicted a very large proportion (73%) of the variance in pure anxiety. Regression analysis #3 presents that IL-16, SCGF, ACEQ4, and sex (all positive correlations), along with CCL3 (negative correlation), predict a substantial part (65.2%) of the variance in SB. Regression #4 shows that IL-16, SCGF, and ACEQ4 (positive correlations) and CCL3 (negative correlation) predict a considerable part (75.2%) of the variance within the phenome. Notably, IL-16 was by far the most robust predictor for all dependent variables mentioned above.

### Results of PLS analysis

**Figure 3** displays the results of a PLS analysis with the phenome of depression, conceptualized as a latent vector extracted from HAMD, HAMA, and SB values, as output variable. Input variables were ACE+NLEs, different CIRS products (e.g. IL-4, sIL-1RA, sIL-2R) and a latent vector extracted from different pro-inflammatory products (the most significant as detected in table 3), including IL-16, IL-18, TRAIL, CCL5, CCL27, HGF, M-CSF, PDGF, SCF, SCGF) (labeled: IRS+GF activation). With an SRMR value of 0.044 the model quality fit was adequate. In addition, the two latent vectors showed adequate convergence and construct validity: all loadings of the phenome factor were ≥0.885 and IRS+GF factor ≥0.683; AVE values were 0.886 and 0.635, respectively; Cronbach’s alpha of both constructs were 0.935; and composite reliability values were 0.959 and 0.945, respectively. Complete PLS analyses performed on 5,000 bootstrap samples showed that 62.3% of the variance in the phenome was explained by the regression on the IRS+GFs factor, IL-4, and ACE+NLEs. There were no significant effects of ACE+NLEs on IL-4. None of the other CIRS components (e.g., sIL-1RA or sIL-2R) was a significant predictor of the phenome beyond the effects of IL-4, which was the most significant CIRS component. Moreover, 27.4% of the variance in IRS+GF was explained by ACE+NLEs. Post hoc analysis (G*Power 3.1.9.4) shows that the achieved power was 0.999. There were no significant effects of age, sex, or BMI on any of the input or output variables. There was a significant specific indirect effect of ACE+NLEs on the phenome that was mediated by IRS+GF (t=5.93, p<0.001).

**Figure 3:**
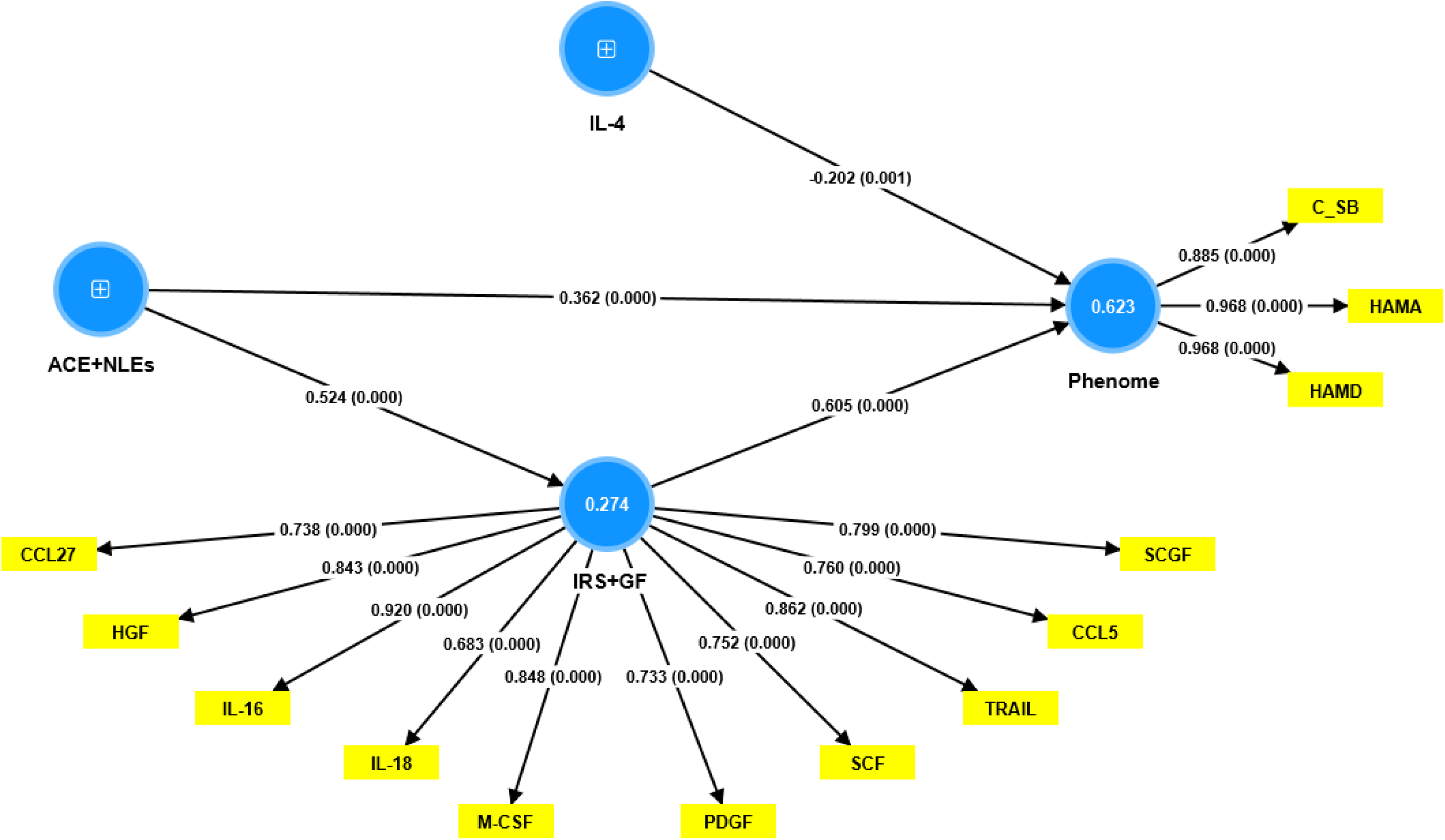
Results of a Partial Least Squares analysis (5,000 bootstraps) with the phenome of depression as output variable, and adverse childhood experiences combined with negative life events (ACE+NLEs) and cytokines, chemokines, and growth factors as explanatory variables. The phenome is conceptualized as a latent vector extracted from the Hamilton Depression (HAMD) and Anxiety (HAMA) rating scales and suicidal behaviors (SB). The immune-inflammatory response and growth factor (IRS+GF) network is conceptualized as a latent vector extracted from the top cytokines, chemokines, growth factors that are induced by ACE+NLEs. IL: interleukins, TRAIL: tumor necrosis factor-related apoptosis-inducing ligand; CCL: C-C motif chemokine ligand; HGF: hepatocyte GF; M-CSF: macrophage colony-stimulating factor; PDGF: platelet derived GF: SCF: stem cell factor; SCGF: stem cell GF. Figures in blue circles denote average variance explained. Shown are the path coefficients (p values) of the inner model, and loadings (p values) of the outer models.

## Discussion

### Effects of ACEs on immune responses

The first major conclusion of the current investigation is that, in contrast to the alternative M2, Th-2, and CIRS immune profiles, ACEs are strongly associated with enhanced classical M1 macrophage, Th-1, IRS, and neurotoxicity immune profiles. Additionally, we discovered a significant correlation between ACEs and Th-1 polarization, IRS-associated cytokines/chemokines (IL-16, IL-18, TRAIL, TNF-a, CCL2, CCL5, CCL27, CXCL9, and CXCL10) and growth factors (M-CSF, SCF, HGF, SCGFB, PDGFB, and SDF1). On the other hand, ACEs had no significant effects on the two main CIRS constituents, IL-4, and IL-10, although there were some weaker correlations between ACEs and markers of T cell activation (sIL-2R) and M2 macrophage activation (sIL-1RA), both of which have CIRS activity.

These results add to those from Maes et al. (2022), who found that elevated ACE levels are linked to increased M1, Th-1, Th-2, Th-17, IRS, and CIRS immunological profiles. However, the latter results were attained utilizing LPS+PHA-stimulated but not unstimulated whole blood from MDMD patients versus controls. In addition, the recent findings add to those of Maes et al. (2022) demonstrating that ACEs significantly increase PGDF. These results indicate that ACEs are accompanied by increased sensitization of IRS and CIRS pathways and growth factors, which are stimulated upon novel immune injuries (Maes et al., 2022). By extension, these heightened immune responses, which are discovered in an in vitro setting (stimulated whole blood), seem to lead to activated inflammatory and growth factor pathways in the in vivo setting, and consequently, in serum.

There is a substantial link between childhood trauma and increased inflammation levels in adulthood, according to a systematic analysis by Baumeister et al. (2016) that analyzed 25 studies (Baumeister, Akhtar et al. 2016). In this context, numerous observational studies have shown that adults who have experienced ACEs frequently display increased levels of inflammation indicators, including C-reactive protein (CRP), and IL-6 (Baumeister, Akhtar et al. 2016, Danese and Baldwin 2017, Moraes, Maes et al. 2017, Flouri, Francesconi et al. 2019, Iob, Lacey et al. 2020). However, Th-1 polarization, IRS activation, enhanced neurotoxicity, and increases in T-cell related cytokines, chemokines, and growth factors are among the numerous other effects of ACEs that the current investigation uncovered.

### Effects of NLEs on the immune system

The second key conclusion of the current study is that, while NLEs have no effect on immune profiles, ACEs have a strong and dramatic impact on immune activation profiles. NLEs did, however, have a significant impact on specific cytokines/chemokines (including IL-16, IL-18, TRAIL, CCL27, and CXCL10) as well as growth factors (such as M-CSF, SCF, HGF, SCGFB, PDGFB, and SDF1). According to studies using ELISA and flow cytometry methods (Maes, Bosmans et al. 1989, Maes, Bosmans et al. 1990, Steptoe, Willemsen et al. 2001, Szabo, Slavish et al. 2020), acute psychological stressors may stimulate the in vitro, stimulated production and serum levels of pro-inflammatory cytokines (such as IL-6, TNF-α, IFN-γ, and IL-1β), as well as the in vivo expression of activation markers (such as CD69+, and HLADR+).

As a result, although there are some differences, ACEs, and NLEs both seem to boost related immune products. For instance, CCL11 (eotaxin) and sIL-2R levels are raised by NLEs rather than ACEs. On the other hand, while ACEs have an impact on TNF-α, CCL2, CCL5, and CXCL9, NLEs have no discernible impact on any of these analytes. The aforementioned observations are intriguing because they demonstrate that depressed individuals can distinguish between the effects of ACEs and NLEs, indicating that bias resulting from information with a negative valence has not played a significant role in the association between ACEs and the depression phenome.

Significantly, our study reveals that the co-occurrence of ACEs and NLEs has an augmented impact on the prediction of classical M1, Th-1, and neurotoxicity immune profiles, surpassing the predictive power of ACEs in isolation. Therefore, it is reasonable to hypothesize that NLEs could potentially enhance immune activation when coupled with the impacts of ACEs. The aforementioned results can be elucidated by the understanding that ACEs lead to a heightened sensitivity of immune functions, and subsequent immune stressors may trigger the reactivation of the sensitized pathways, as posited by Maes et al. (2022).

However, the differentiation between the impacts of ACEs and NLEs is impeded due to the robust correlation between ACEs and present NLEs. The aforementioned phenomenon can be elucidated by the hypothesis that individuals who have experienced ACEs are more likely to encounter a greater number of NLEs (Kendler, Karkowski et al. 1999). The study conducted by Lob et al. revealed a noteworthy finding that the association between ACEs during the later stages of childhood or adolescence and depressive symptoms was more robust compared to the earlier ACEs (Iob, Lacey et al. 2022). Therefore, depressed individuals exhibit differential discernment not only between ACEs and NLEs, but also between ACEs that occurred earlier or later in life.

### IRS activation mediates, in part, the effects of ACEs on the depression phenome

Multiple studies have reported a significant association between ACEs and the occurrence of recurrent depression and suicidal ideation later in life (Cheong, Carol et al. 2017, Moraes, Maes et al. 2017, Tsehay, Necho et al. 2020, Giano, Ernst et al. 2021, Satinsky, Kakuhikire et al. 2021, Iob, Lacey et al. 2022). Extensive research has been conducted on the correlation between ACEs, such as child abuse, parental separation, and parental mental health issues, and a wide range of negative health outcomes, including MDD and MDMD (Bellis, Hughes et al. 2019, Maes, Rachayon et al. 2022). Previous studies have indicated that ACEs are associated with lifetime suicidal behaviors, illness recurrence, and the severity of the depression phenome, as measured as a latent vector derived from the severity of depression, anxiety, and current suicidal behaviors (Maes, Congio et al. 2018, Maes, Moraes et al. 2019, Maes, Moraes et al. 2021, Maes, Rachayon et al. 2022). Research has demonstrated that acute episodes of depression are linked to recent NLEs (Paykel, Emms et al. 1980, Kendler, Karkowski et al. 1999, Kraaij, Arensman et al. 2002). Consistent with this existing literature, our findings indicate that the confluence of NLEs and ACEs is a significant predictor of the intensity of the depression phenome.

The present study’s third primary discovery indicates that the severity of depression/anxiety symptoms, suicidal behaviors, and the phenome of FE-MDMD are significantly predicted by the effects of immune activation, growth factors and ACE+NLEs. Moreover, the findings from our study, as demonstrated through the PLS analysis, suggest that the activation of the IRS and growth factors serves as a partial mediator for the impact of ACE+NLEs on the phenome. The research conducted by Maes, and colleagues (2022) also indicated that immune stimulation and enhanced growth factor levels play a partial mediating role in the impact of ACEs on the phenome of FE-MDMD. This implies that there are additional pathways that contribute to this relationship. It is noteworthy that the impact of ACEs on suicidal tendencies and depression is influenced by numerous immune-related mechanisms, such as alterations in the microbiome and a distinct gut dysbiosis enterotype, oxidative stress leading to lipid peroxidation and protein damage, decreased antioxidant defenses, and reduced levels of nerve growth factor (Maes, Moraes et al. 2019, Maes, Moraes et al. 2021, Maes, Rachayon et al. 2023, Maes, Vasupanrajit et al. 2023).

### Importance for the pathophysiology of MDD and MDMD

Prior research has revealed that severe MDD or MDMD is characterized by crucial processes such as IRS activation and heightened neurotoxicity. Additionally, Th-1 activation is believed to be one of the primary processes involved (Maes, Bosmans et al. 1990, Maes 2011, Almulla, Ali Abbas Abo et al. 2023, Rachayon, Jirakran et al. 2023). MDMD is distinguished by a rise in the count of activated effector T cells, namely CD4+CD71+, CD4+CD40L+, CD4+CDHLADR+, as well as cytotoxic T cells, namely CD8+CDL40+, and an increase in T cell activation-driven proliferation of B cell phenotypes that stimulate IgM-mediated autoimmune processes (Rachayon, Jirakran et al. 2023). Furthermore, the current research demonstrates that a deficiency in IL-4, a crucial cytokine in the context of CIRS functions, exacerbates the effects of IRS, T cell activation, and heightened neurotoxicity on the depression phenome. Phrased differently, the immune stimulatory effects of ACEs will result in a stronger impact in subjects with lowered IL-4 production.

In addition, MDD is associated with reduced protection from T regulatory cells, specifically those expressing CD25+FoxP3+GARP+ and CD25+FoxP3+CD152+ (Maes, Rachayon et al. 2023, Rachayon, Jirakran et al. 2023). Our findings suggest that the breakdown of immune tolerance is an additional factor that increases susceptibility to the neurotoxic effects of IRS activation. Heightened levels of IL-16 may play a crucial role in this sequence of events (Almulla, Ali Abbas Abo et al. 2023). This is because IL-16 binds to CD4, which subsequently triggers T cell activation and chemotaxis (Mathy, Scheuer et al. 2000, Skundric, Cruikshank et al. 2015, Gregory 2021).

Overall, our research indicates that ACEs and the combination of ACEs and NLEs affect the phenome, and that these effects are partially mediated by T cell activation, Th-1 polarization, and increased T cell-associated cytokines, chemokines, and growth factors, which have a strong effect. Notably, the neuroimmunotoxic effects of ACE+NLEs in MDMD may exacerbate the neurotoxic effects of elevated oxidative stress toxicity, coupled with diminished antioxidant or neurotrophic defenses, and the effects of gut dysbiosis (Maes, Moraes et al. 2019, Maes, Moraes et al. 2021, Simeonova, Stoyanov et al. 2021, Maes 2022).

## Limitations

Our study has limitations that must be considered when interpreting the results. Given that the immune-inflammatory response is always intertwined with oxidative and nitrosative stress, it would have been more intriguing to evaluate biomarkers for oxidative and nitrosative stress in FE-MDMD patients. Future research should investigate the effect of ACEs and NLEs on the reverse cholesterol transport pathway. In this context, we recently discovered that affective disorders are accompanied by decreased reverse cholesterol transport, a robust antioxidant and anti-inflammatory defense mechanism (Almulla, Thipakorn et al. 2023).

## Conclusions

The present study shows that the first onset of depression is significantly associated with elevated levels of ACEs and NLEs. Moreover, the impact of ACEs and NLEs on FE-MDMD is partially mediated by an increased immune-inflammatory and growth factor responses. Hence, a potential proactive strategy for deprogramming the effects of ACEs and mitigating the influence of NLEs on IRS activation and the growth factor network could target the heightened growth factor and immune responses, especially T cell activation, as well as enhancing immune tolerance by stimulating CIRS and Treg cell functions.

## Data Availability

The data compiled and evaluated throughout this study will be accessible through the corresponding author (M.M.), provided a reasonable request is made. Please note that data availability will be ensured once the authors fully utilise it for their intended research purposes.

## Acknowledgments

The authors thank everyone at Al-Hakeem General Hospital who contributed to the data collection.

## Ethical approval and consent to participate

This study received approval from the Ethics Committee of the College of Medical Technology at the Islamic University of Najaf, Iraq (Document No. 18/2021). The research was conducted strictly following local Iraqi and international ethical and privacy regulations. This includes but is not limited to principles outlined in the World Medical Association’s Declaration of Helsinki, the Belmont Report, the CIOMS Guideline, and the International Conference on Harmonization of Good Clinical Practice. Our institutional review board is committed to maintaining the highest standards, strictly adhering to the International Guideline for Human Research Safety (ICH-GCP).

## Author’s contributions

Patient-related tasks and blood sample collection were managed by AFA and AAA. AFA and MM were responsible for the measurement of serum biomarkers. The statistical analysis was conducted by MM. Both AFA and MM contributed to the manuscript’s composition and editing. All authors participated in reviewing and approving the definitive version of the manuscript.

